# Causal Factors of Effective Psychosocial Outcomes in Online Mental Health Communities

**DOI:** 10.1101/2020.08.15.20175836

**Authors:** Koustuv Saha, Amit Sharma

## Abstract

Online mental health communities enable people to seek and provide support, and growing evidence shows the efficacy of community participation to cope with mental health distress. However, what factors of peer support lead to favorable psychosocial outcomes for individuals is less clear. Using a dataset of over 300K posts by *∼*39K individuals on an online community TalkLife, we present a study to investigate the effect of several factors, such as adaptability, diversity, immediacy, and the nature of support. Unlike typical causal studies that focus on the effect of each treatment, we focus on the outcome and address the *reverse* causal question of identifying treatments that may have led to the outcome, drawing on case-control studies in epidemiology. Specifically, we define the outcome as an aggregate of affective, behavioral, and cognitive psychosocial change and identify *Case* (most improved) and *Control* (least improved) cohorts of individuals. Considering responses from peers as treatments, we evaluate the differences in the responses received by *Case* and *Control*, per matched clusters of similar individuals. We find that effective support includes complex language factors such as diversity, adaptability, and style, but simple indicators such as quantity and immediacy are not causally relevant. Our work bears methodological and design implications for online mental health platforms, and has the potential to guide suggestive interventions for peer supporters on these platforms.

## 1 Introduction

Online Mental Health Communities (OMHCs) are dedicated online support platforms aimed at aiding individuals to share, discuss and solicit information and support related to mental health. In many ways, OMHCs function like the online analog of support groups (Potts 2005). Anonymity and social connectedness in OMHCs help individuals overcome stigma and make candid self-disclosures about their mental health concerns (Andalibi et al. 2016). Examples of OMHCs include mental health subreddits on Reddit, condition-specific discussion forums on 7Cups, and social network-based interactions on Talklife (Pruksachatkun et al. 2019). OMHCs help individuals draw psychosocial benefits that help them cope with their mental health struggles (Love et al. 2012).

Given the growing popularity of OMHCs, research has studied various aspects of participation in these communities and how they may lead to better psychosocial outcomes for individuals. Extending evidence from experiments that demonstrate the efficacy of online support (Winzelberg et al. 2003), studies on Reddit and TalkLife find that they offer a thriving, global community for people to talk about their mental health (Pendse et al. 2019). They provide a fine-grained data source to understand how people express mental health distress and support each other in the real world, such as shifts in suicidal ideation (De Choudhury et al. 2016).

However, relatively little attention has been directed on the peer supporters on such platforms and how they can be more effective at providing support. A natural question to ask is *what kinds of supportive behavior leads to better outcomes for individuals receiving support*. Identifying support characteristics in responses and discussions that lead to positive psychosocial outcomes can yield insights on the best strategies of providing support, complementing work in psychotherapy literature (Norcross and Lambert 2018). Further, by focusing on natural conversations *in situ*, these insights can help OMHC owners design recommendations for their members to make more effective supportive responses.

To investigate the factors that contribute to effective support, we adopt the “case-control” study design from epidemiology (Schulz and Grimes 2002). The idea is to identify individuals who have had positive outcomes (*Case* group) and then retrospectively compare their characteristics with a similar *Control* group of individuals. Specifically, we identify people who have had long-term positive psychosocial changes and compare the characteristics of responses they received to that received by those who did not have such positive changes. In the language of causal inference, each response from a peer supporter can be considered as an intervention for an individual, and we are interested to find the characteristics of interventions that lead to the maximum positive change. Using the symmetry of the back-door method (Pearl 2009), we argue that looking for interventions that vary significantly between case and control groups translate to finding interventions with causal effects on the outcome.

Specifically, we work with a longitudinal dataset of *∼*39K individuals on TalkLife, an online mental health platform. We quantify their psychosocial outcomes as an aggregate measure of affective, behavioral, and cognitive outcomes. On the basis of psychosocial change from when they joined to the present, we obtain two separate cohorts of individuals — psychosocially most (*Case*) and least (*Control*) improved. We compare the differences in responses across a range of characteristics drawn on psychotherapy literature like adaptability, immediacy, diversity, emotionality, language style, and nature of support. Confirming past work, we find complex linguistic attributes such as adaptability, diversity, and style are significant factors for driving positive psychosocial change. In particular, factors related to adaptability such as topical congruence and linguistic accommodation have the highest difference between *Case* and *Control* groups. Somewhat surprisingly, the average length of responses has a substantial positive effect towards driving people to better outcomes, possibly as a proxy for the linguistic factors described above. Other simple factors, however, such as number of responses and immediacy of receiving a response do not have significant differences between *Case* and *Control*.

Compared to previous work by De Choudhury and Kıcıman (2017) estimating effects of using specific phrases in responses, our work has an advantage whenever one is interested in analyzing continuous treatments and *finding* the interventions that lead to desired outcomes. This is because most *forward* causal inference methods (Gelman and Imbens 2013) require binarization of treatment variables. In contrast, case-control methods avoid apriori binarization of complex treatments and estimate the differences in treatment instead. Especially in OMHCs where everyone typically receive responses, our proposed method is useful to determine the necessary *dosage* increase of support treatments that can increase the likelihood of positive outcomes (Hernan and Robins 2010). There is also a computational advantage. In forward causal inference methods (Rubin 2005), one may estimate a separate propensity score model for each treatment whereas our case-control method allows estimating the differences in multiple treatments at once. We discuss the methodological and practical implications of our work towards improving support in OMHCs through recommendation-based interventions for peer supporters.

### Privacy, Ethics, and Disclosure

This paper uses sourced data (licensed and consented) from TalkLife. Our work is in collaboration with TalkLife, and given the sensitivity of our work, we are committed to securing the privacy of the individuals. The dataset was accessed through secured databases with necessary privacy and ethical protocols in place, and the dataset was de-identified and no personally identifiable information was used. This paper only reports aggregated measures to prevent traceability and identifiability of individuals on the platform. Even accounting for the benefits, we recognize the potential misuses, risks, and ethical consequences involved with this kind of research, which we elaborate in Discussion. This work is approved by the Institutional Review Board at Microsoft Research.

## 2 Background and Related Work

### Effective Psychotherapeutic Interventions

What constitutes effective counseling and psychotherapeutic strategies has interested researchers and practitioners for a long-term now (Labov and Fanshel 1977), and treatments and therapeutic methods constantly evolve and advance over time. Lambert and Barley (2001) formulated four areas that influence a care-seekers’ outcome in psychotherapeutic settings – extratherapeutic factors, expectancy effects, specific therapy techniques, and common factors. Among these, common factors include empathy, warmth, congruence, therapeutic alliance, are found to be most highly correlated with the outcomes. In another work, Norcross and Lambert (2018) conducted a meta-analysis on the effectiveness of several elements of psychotherapeutic relationships.

In the area of online technology aided and mediated mental health interventions, Cavanagh et al. (2018) showed the efficacy of computer-mediated psychotherapy towards positive clinical outcome. Although its efficacy is yet to be established and results are mixed (Rollman et al. 2018), researchers have stressed the importance of social media as a mental health intervention platform (Chikersal et al. 2020, Ernala et al. 2017, Merolli et al. 2013, Yoo and De Choudhury 2019). Relatedly, Dinakar et al. (2015) used computational linguistics and machine learning to improve crisis counseling and interventions, Haberstroh et al. (2007) studied online counseling experiences, and Althoff et al. (2016) studied effectiveness of counseling language including adaptability, creativity, and perspective change. Our work draws upon these prior works to evaluate what factors help in desirable psychosocial outcomes in “online psychotherapeutic setting”, where individuals seek and share mental health related support.

### Support in Online Mental Health Communities (OMHCs)

With the widespread use of social media-based technologies, OMHCs are becoming increasingly popular. Originally ideated as online analogs of support groups, individuals in these communities share and seek support related to sensitive mental health concerns faced by themselves or their loved ones (De Choudhury and De 2014; Huh 2015; Saha et al. 2019a; 2020; Sharma and De Choudhury 2018; Kummervold et al. 2002). Prior work studied how anonymity, engagement, social capital, and social connectedness help in candid self-disclosure and seeking mental health support (Andalibi et al. 2016; Ernala et al. 2018). In addition, online social support is known to build interpersonal relationships, and to improve psychological wellbeing, self-esteem, satisfaction, and reciprocity (Steinfield et al. 2008; Oh et al. 2013).

For specialized OMHC platforms such as Talklife or 7Cups, recent studies have examined positive outcomes over a sample of users (Baumel et al. 2018) or over a single thread or bursts of conversation (Kushner and Sharma 2020, Pruksachatkun et al. 2019; Pendse et al. 2019). We extend this research by focusing on long-term changes in one’s mental health over a large sample of individuals and retrospectively finding the most relevant causes.

### Causal Inference Studies on Observational Data

The gold-standard approach to establish causality is via a randomized controlled trial. In early work, such trials were conducted to assess the efficacy of online support communities of breast cancer (Winzelberg et al. 2003). However, trials are not always feasible due to practical and ethical concerns (Hannan 2008). As an alternative, researchers resort to observational studies. While these cannot guarantee causality, observational studies allow to investigate long-term and longitudinal data, and are especially useful to find candidate treatments for a future randomized trial when no preferred treatment is known apriori (Rubin 2005). There are two popular means of conducting observational studies — 1) cohort based, where the treatment is known and the goal is to find its causal effects on the outcome, and 2) case-control based, where the outcomes are known, and the goal is to (retrospectively) find the treatments that potentially caused the outcome.

In our related space of social media and mental health, observational studies have examined the effects of suicidal ideation (De Choudhury et al. 2016), social support (De Choudhury and Kıcıman 2017), psychiatric medications (Saha et al. 2019b), exercise (Dos Reis and Culotta 2015), alcohol use (Kıcıman et al. 2018), crisis (Saha and De Choudhury 2017), and counseling interventions (Saha et al. 2018). These studies adopted the cohort based, or prospective analysis of conditioning on treatments, and matching on similar individuals to examine the differences in the outcomes, which finally quantifies the causal effects (Olteanu et al. 2017). However, cohort-based studies may not be suitable when the goal is to rank multiple continuous treatments on their causal effect, especially when almost everyone receives variable treatment dosage, and there are no obvious way to binarize the different treatments. We therefore adopt the case-control study design, and provide a method to analyze the effects of several continuous treatments.

## 3 Data

The dataset of our study comes from TalkLife (Pendse et al. 2019), an online mental health discussion forum, self-describing itself as a “safe social network to get help and give help”. From the standpoint of social computing interface, TalkLife functions like many other online communities and discussion forums. The community members participate via discussion threads, where each discussion thread consists of an original post (or “post” hereon), and a number of responses by community members which are typically relevant to the post in the discussion thread. We obtain data from the TalkLife platform (in collaboration with TalkLife) from August 2011 to January 2019. This data includes discussion threads, each with a single post and a number of responses by community members. We note that TalkLife responses can also follow a hierarchical nature, however our work accounts for all kinds of responses similarly, under an umbrella term of “responses”. Our dataset consists of a total of over 6.5M posts (equal number of discussion threads) and 20M responses posted by over 300K users. On average, each discussion thread receives 4.44 responses (stdev. = 30.60). Table 1 shows examples of four paraphrased posts on TalkLife.

**Table 1:**
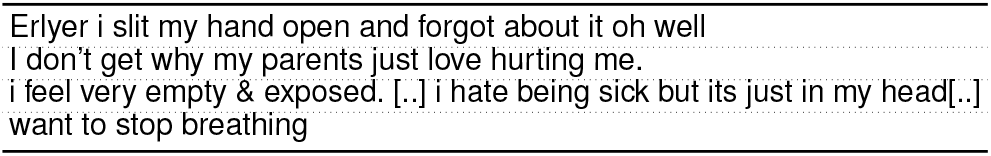
Example paraphrased posts on TalkLife.

Social computing platforms tend to change significantly over the years, which could also lead to changes in usage behavior and objectives of individuals. For the purposes of our study, to minimize the effects of long-term platform-level changes and aggregated use behavior, we limit our analysis to 38,977 individuals who started participating in TalkLife after January 01, 2016, continued participation (posted more than once) beyond January 01, 2018, and overall had posted at least 15 times on the platform. Given that our data lasts till January 2019, each user has been on the platform for at most three years (mean = 222 days, median = 142 days). Given that the TalkLife platform has stabilized over recent years, we expect a lower impact of platform-level changes during this three-year time period. This study concerns a dataset comprising of 3,184,612 posts and 23,528,159 responses with 4.72 response per post (stdev. = 54.81). Figure 1 shows the distribution in our dataset as responses per post (Figure 1(a)), posts per user (Figure 1(b)), and responses per user (Figure 1(c)).

**Figure 1:**
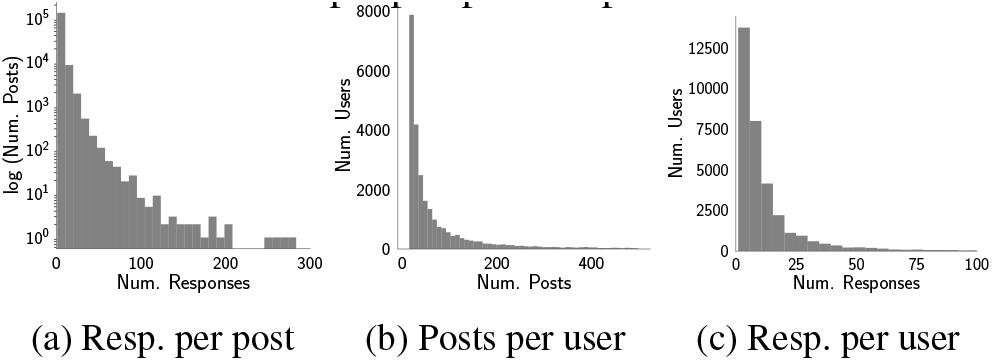
Distribution of posts and responses in study dataset.

## 4 Study Design and Methods

### Retrospective Case-Control Design

We are interested in the effect of different kinds of supportive interventions (or treatments) on psychosocial outcomes for users on OMHCs. If interventions are well-defined (e.g., in medicine, whether a drug was prescribed), then the most common approach is to estimate the effect of each intervention separately. If *T* is the treatment, *Y* the outcome, and *W* represents common causes or confounders of *T* and *Y*, then the causal effect (Pearl 2009) of *T* on *Y* is represented as:

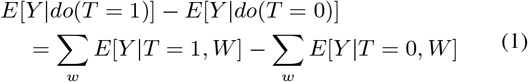

Effectively, the method compares outcomes for people with or without the intervention while conditioning on all confounders. However, when there are multiple candidate treatments for the same outcome of interest, it may be more appropriate to ask the *reverse* causal question. That is, rather than the effect of a treatment, we ask about the potential causes of an observed outcome. In our problem, for example, the outcome is pre-specified—psychosocial health of individuals—but treatments are not. At one level, we could consider participation on Talklife as a means to be treated. At another level, we would like to consider the effect of several characteristics of language and behavior used by peer supporters and isolate their effects. Thus, our goal is to *find* treatments that lead to a significant change in the outcome.

When the outcome is well-specified, and it distinguishes two groups of *Case* and *Control* users, we can use the following estimator for finding the interventions that causally affect the outcome, based on the case-control study design in epidemiology (Schulz and Grimes 2002):

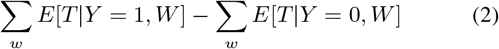

In our work, *W* corresponds to covariates consisting of individual attributes. Intuitively, this equation refers to conditioning on *W*, and then calculating the differences between treatment for *Case* and *Control*. The estimator compares individuals with a positive outcome (*Y* = 1) with others (Y = 0) and measures the difference in intervention values between the two groups, while conditioning on all known confounders. This is the so-called ‘reverse” causal inference problem (Gelman and Imbens 2013). Whenever there is a significant change in *T* from *Y* = 0 to *Y* = 1 keeping all confounders *W* constant, it implies that there is a causal effect of *T* on *Y*. Given the same outcome, we can do this analysis repeatedly for finding the treatments with the highest effects. As we will see later, the case-control analysis provides some computational benefits especially when working with continuous intervention variables and selecting a suitable cutoff to binarize them for a future randomized experiment. Further, in contrast to cohort-based causal inference studies that condition on individual treatments, our approach allows accounting for a combination of multiple treatments together on the same individuals (emulating closer to real-world settings).

The rest of this section contextualizes the above estimator in an OMHC. We operationalize the outcome on psychosocial health, determine the case and control groups, and then finally list potential interventions that we test.

### Measuring the Outcome: Psychosocial Health

Towards our research objective of understanding effective psychotherapeutic interventions, we first operationalize “improvement in mental health outcome” as observed on Talk-Life. Researchers have argued on what constitutes improvement and success in psychotherapeutic, psychological, and psychiatric care (Perkins 2001), where traditionally symptom reduction has been considered to be the improvement in quality of life. Generally speaking, “psychosocial health” is considered as an appropriate terminology that not only encompasses both psychological and social wellbeing, but also places the locus of health in the individual by including social wellbeing in the form of social adjustment and environmental response (Larson 1996). We situate our work on the impacts of social media based interventions on one’s psychosocial health and wellbeing (Merolli et al. 2013). Given that psychosocial health is a complex construct and there is no easy means to quantify it, we adopt a conservative definition of psychosocial health based on observed behavior on the platform. As a user’s posting behavior is our only available data, we draw upon prior work that operationalize therapeutic responses in online mental health communities and social media, grounded on psychology, psychiatry, and expressive writing literature (Ernala et al. 2017, Saha et al. 2018). We broadly group these observed psychosocial outcomes in three categories — *affective, behavioral*, and *cognitive* outcomes (Breckler 1984), and then aggregate them to construct a single outcome metric.

#### Affective Outcomes

Simplistically, affect refers to an emotional response, and affective behavior is indicative of one’s psychological wellbeing. Because social media posts are written in a self-motivated and self-initiated fashion, language is a strong means to infer affective psychosocial health. To measure this, we use the following:

##### Affective Words

We use the psycholinguistic lexicon, Linguistic Inquiry and Word Count (LIWC) (Pennebaker et al. 2003) to obtain proportion of affective (positive and negative affect) keywords per user. This draws upon expressive writing literature which associate language with therapeutic symptoms. Increased use of positive affect and decreased use of negative affect words correspond with psychosocial improvement.

##### Language Indicative of Mental Health Symptomatic Outcomes

To identify the presence of mental health concerns, prior work built machine learning classifiers of social media language indicative of depression, anxiety, stress, suicidal ideation, and psychosis (Saha et al. 2019b). These are *n*-gram (*n* = 1,2,3) based binary SVM models. For training these classifiers, the positive class comes from domain dependent data on Reddit (*r/depression, r/anxiety, r/stress, r/SuicideWatch*, and *r/psychosis* subreddits for the corresponding classifiers), and the negative training examples come from a set of random non-mental health related content from Reddit. Similar to Saha et al. (2017; 2019b), we conduct linguistic equivalence test — cosine similarity of the word embedding representations of top 500 *n*-grams in the Reddit and Talklife datasets shows a high similarity of 0.92. This entails very similar transfer datasets, and similar performance, given that the classifiers have performed reasonably well when transferred on other social media datasets (Saha et al. 2019b). Using these classifiers, we obtain the aggregated proportion of posts that express mental health concerns corresponding to each Talk-Life user. That is, lower the proportion of posts expressing mental health concerns, better is one’s psychosocial health.

#### Behavioral Outcomes

Behavioral psychosocial health consists of an individual’s overt actions, behavioral intentions, and verbal statements regarding behavior (Breckler 1984). Behaviors such as changes in social functioning and shift of interests could be indicative of an individual’s changing psychological trajectory (Saha et al. 2018, Guntuku et al. 2019). To quantify behavioral psychosocial outcomes, we obtain three attributes on an individual’s behavior on the platform. The first of these is *activity*, or the frequency of participation on the platform – this is quantified as the number of posts per day for every individual. The second is *interactivity*, or how interactive an individual is — this is quantified as the ratio of the number of responses (to others’ posts) to the number of self-posts. This essentially quantifies an individual’s behavior of providing support compared to seeking support. The final one is *interaction diversity*, or the topical diversity of discussions an individual engages in — each discussion thread is labeled with a particular topic (eg., relationships, family, self-harm, friends, hopes, etc.) by the original poster. These measures are directly associated with psychosocial health — an increase in these measures corresponds to an improvement in psychosocial health (Saha et al. 2018).

#### Cognitive Outcomes

Beliefs, knowledge structures, perceptual responses, and thoughts constitute cognitive component of psychological wellbeing (Breckler 1984). Cognitive attributes is another indicator of an individual’s psychological health (Bandura 1993). Drawing on psycholinguistics literature that demonstrates how the style and structure in language define one’s cognitive behavior, we adopt the following measures to define cognitive psychosocial health.

*Readability* measures the ease with which a reader can understand a given text. We adopt the Coleman-Liau Index (CLI) which provides readability assessment based on character and word structure within a sentence, calculated as, *CLI* = (0.0588*L −* 0.296*S −* 15.8), where *L* is the average number of letters per 100 words, and *S* is the average number of sentences per 100 words. A greater CLI measure indicates a better writing quality, and an increase of CLI indicates psychosocial improvement (Ernala et al. 2017).

*Complexity and Repeatability* capture one’s cognitive state in the form of planning, execution, and memory (Ernala et al. 2017). We quantify complexity as the average length of words per sentence, and repeatability as the normalized occurrence of non-unique words. While linguistic complexity has a positive association with one’s psychosocial health, repeatability shares a negative association with the same.

##### Psycholinguistic Keywords

We use LIWC lexicon to obtain the proportion of keywords corresponding to cognition, perception, and linguistic style categories, where linguistic style keywords correspond to non-content keywords in language such as, temporal references (past, present, and future tense), lexical density and awareness (auxilliary verbs, preposition, adverbs, verbs, articles, conjunctions, inclusive, and exclusive), and interpersonal focus (1st person singular and plural, 2nd person, and 3rd person pronouns). Literature posits the importance of keywords in understanding cognitive behavior. For instance, the variations in pronoun use reflects the transformation in the way individuals think about themselves with respect to others, and the use of articles and adverbs could indicate how individuals process complex narratives (Pennebaker et al. 2003). A greater use of these keywords is associated with one’s improved psychosocial state.

##### Overall Psychosocial Outcome

After normalizing each of the above outcomes on a min-max scale of 0 to 1, we operationalize psychosocial health of an individual as a composite measure of unit-weighted and sign-adjusted average across each of the outcomes so that higher values indicate a better psychosocial health (see below). As noted before, while this cannot be argued to be perfect, we believe that by accounting for several symptomatic observable changes on the platform, such a composite measure should be theoretically correlated with the actual psychosocial health of an individual.

> *outcome = μ(pa–na–mh_language+activity+int._diversity+interactivity+readability–complexity–repeatab.+cog._words)*

### Determining *Case* and *Control* Individuals

To understand the effects of support, social interactions, and responses (treatment), we identify and distinguish those individuals who improved the most, and those who did not after a period of time on the platform. Adopting terminologies from epidemiological observational studies, we name these groups as *Case* (improved) and *Control* (not improved or worsened) (Schulz and Grimes 2002). Ideally the improvement should be determined based on the change in mental health state in the present from their initial state on the platform (or before they are treated). As a proxy of the initial state, we quantify an individual’s baseline psychosocial health on their first *n*_1_ posts since they joined the platform. Treatment correspond to the attributes of responses received on the next *n*_2_ posts on the platform and the outcome as the average psychosocial health over all posts after *n*_2_. Essentially, we draw on variable treatment effect framework (Hernan and Robins 2010), and segregate an individual’s timeline of activities on the platform into pre-treatment phase, treatment phase, and post-treatment phase (see Figure 2 for a schematic overview of the segregation on an individual’s timeline). We choose the number of posts by a user instead of duration on the platform since it provides a better metric for exposure to responses given high variance in people’s activity.

**Figure 2:**
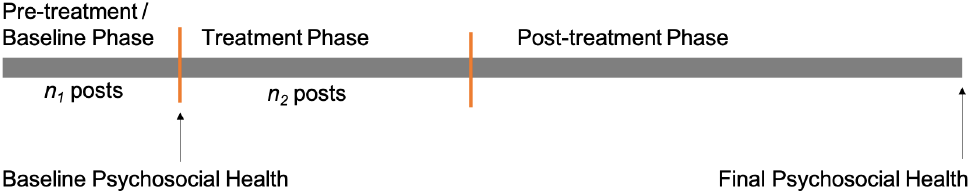
Schematic figure of a user’s timeline in our study design. Psychosocial health is determined as an average of observed measures in the corresponding period.

**Figure 3:**
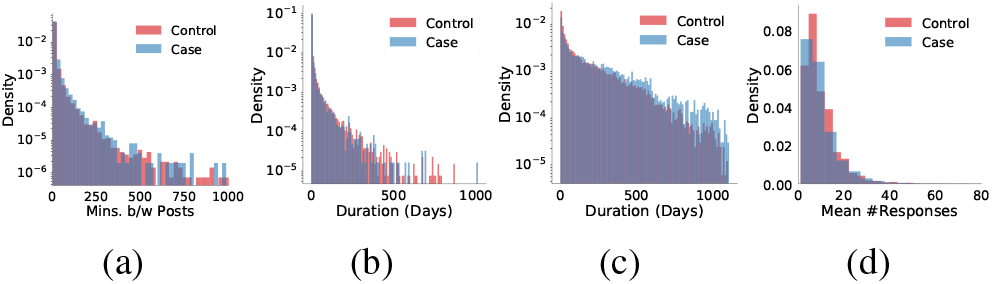
Distribution in *Case* and *Control* in terms of (a) time between successive posts (b) time period in treatment phase, (c) time period in post-treatment phase, (d) number of responses (per user) received in the treatment phase.

#### Choice of *n*_1_ and *n*_2_

There is a tradeoff in choosing *n*_1_. A smaller *n*_1_ ensures that we capture the initial state of a user without the effects of responses, but also exposes us to high variance in estimating it. We thus vary combinations of *n*_1_ and *n*_2_ in different values between 2 and 8 posts, and check for consistency in our findings. We observe that our results are not sensitive to the choice of *n*_1_ and *n*_2_. For the ease of exposition, we first discuss and report findings with pretreatment phase of *n*_1_ = 3 and treatment phase of responses received in the next *n*_2_ = 8 posts per individual. Following this, we revisit the robustness of our findings for different combinations of *n*_1_ and *n*_2_.

#### Case and Control Individuals

Within our dataset, we find that psychosocial change ranges between −0.27 and 0.36, with a mean change of 0.04 (std. = 0.05). For the purposes of our study, we define *Case* to be the individuals who lie in the top 80 percentile of psychosocial change, and *Control* to be the individuals in the bottom 50 percentile of psychosocial improvement (see Figure 4a). Our choice of 80% is motivated by the idea of restricting *Case* to only people with *very good* outcomes so that we can better understand the nature of support interventions behind those changes. With this definition, we obtain 6,789 *Case* and 16,972 *Control* individuals, whom we study for our ensuing analyses.

**Figure 4:**
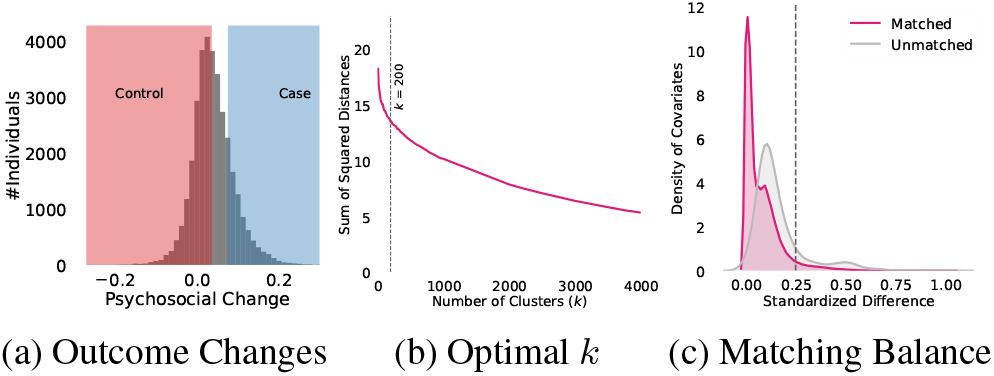
(a) Dist. of Psychosocial outcome of all individuals, (b) *k*-means clustering of individuals on covariates for several *k*, (c) Standardized differences following matching.

#### Testing Comparability of *Case* and *Control*

Before conducting causal analysis on *Case* and *Control*, we evaluate if their data is comparable. We compare the duration between successive posts made by *Case* and *Control* individuals (Figure 3a). We find that *Case* posts are separated by an average 20 minutes and *Control* posts are separated by an average 18 minutes. There is no statistical significance as per independent sample *t*-test and Kolmogorov-Smirnov (*KS*) test (*p>*0.1). Given that our study design rests upon a threshold on the number of posts for specifying the treatment phase (Figure 2), we test if this specification leads to any biases in length of participation time period by comparing the distribution of time period per *Case* and *Control* individual in treatment (Figure 3b) and post-treatment (Figure 3c) phases. The mean length of time in the treatment-phase is 15 days for *Case* and 16 days for *Control* individuals, and the same in the post-treatment phase is 221 days for *Case* and 204 days for *Control* individuals. For both comparisons we find no statistical significance as per *t*-test and *KS*-test (*p>*0.1). Again, because we consider receiving responses as treatment, we compare if *Case* and *Control* received different number of responses overall, where we find no statistical significance as per *t*-test and *KS*-test (*p>*0.1). These tests provide evidence that *Case* and *Control* datasets are comparable, with minimal biases due to unaccounted measures.

### Matching of Similar Individuals

We next aim to *identify the causes* of post-treatment psychosocial outcomes. We adopt a case-control framework, that conditions on the outcomes to differentiate the treatment between *Case* and *Control*. Theoretically, given two similar individuals, their likelihood to improve is similar if they were subjected to the same treatment. Thus, outcome difference is potentially caused by the differences in treatment, provided the biases due to confounders are minimized (Silber et al. 2001). We assume that all potential interventions are via responses on TalkLife, or more generally that interventions outside the platform similarly affect both *Case* and *Control*.

#### Covariates

To reduce biases associated with confounders, the first step involves identifying a suitable set of covariates. The covariates include the exact same affective, behavioral and cognitive measures of psychosocial health, as described in the previous subsection on outcomes. However, while the outcomes are measured over posts that come after the first *n*_1_ + *n*_2_ posts, the distinction is that we compute these covariates for matching using only their first *n*_1_ posts. In effect, we control for covariates that are baseline behavioral and psychological attributes of individuals. For each covariate, we quantify an aggregated measure per individual within their pre-treatment (first *n*_1_) posts and responses received to them. The covariates include their pre-treatment psychosocial measures, which are: affective measures (normalized quantity of affective words and classifiers of depression, anxiety, stress, psychosis, and suicidal ideation), behavioral measures (activity, interactivity, and interaction diversity), and cognitive measures (readability, complexity, and repeatability). The covariates additionally include the top 500 *n*-grams (*n* = 1,2,3) per user, and the pre-treatment average number of responses received per post per individual. The choice of covariates is motivated by prior work (Kıcıman et al. 2018, Saha et al. 2019b). We use these covariates as features in clustering similar users in our ensuing matching step.

#### Matching Approach

To find statistically comparable individuals, we use matching. This simulates a randomized trial setting by conditioning on as many as covariates as possible (Rubin 2005). To compare against counterfactual scenarios, for those who improved (*Case*) we find their similar (matched) counterparts among the ones who did not improve (*Control*). Typically, matching methods match individuals on the basis of the likelihood of being treated, however, in our case every individual is treated (or exposed to responses), although the “dosage” of treatment measures may vary. To account for variable treatment across individuals (Hernan and Robins 2010), we match individuals in an unsupervised fashion using *k*-means clustering. This approach functions like a stratified matching approach (Kıcıman et al. 2018), where each cluster (or stratum) consist of matched individuals.

To determine the number of clusters (*k*) in *k*-means, we use the well-adopted elbow heuristic — optimal *k* can be located around the greatest drop in density across clusters. Figure 4b plots the sum of squared distances of samples to the nearest cluster centroids for *k* varying between 1 and 5,000. Manually inspecting Figure 4b and using Kneedle algorithm (Satopaa et al. 2011), we approximate that the greatest drop (maximum curvature) occurs at around *k* = 200, which we adopt as the number of clusters in our analysis.

After we cluster similar individuals, we drop those clusters without sufficient number of *Case* and *Control* users as these clusters could lead to biased findings (Kıcıman et al. 2018). Using a threshold of at least 10 *Case* and 10 *Control* user per cluster, we obtain 181 usable clusters that contain 6,758 *Case* and 16,920 *Control* individuals in total — together 99.6% of the Case-Control users that we identified. Each cluster essentially contains similar *Case* and *Control* individuals conditioned on pre-treatment covariates (psychosocial health and language on the platform). For better interpretation, we label the clusters by ranking on average interactivity of cluster members, i.e., those with greater interactivity are more likely to be placed at a higher value cluster. For a sample of interpretable covariates, Figure 5 shows differences between the clusters on multiple dimensions — for e.g., consider the pair of first two clusters, while the first cluster shows higher anxiety and higher interaction diversity, the second cluster shows higher suicidal ideation, anger, and activity.

**Figure 5:**
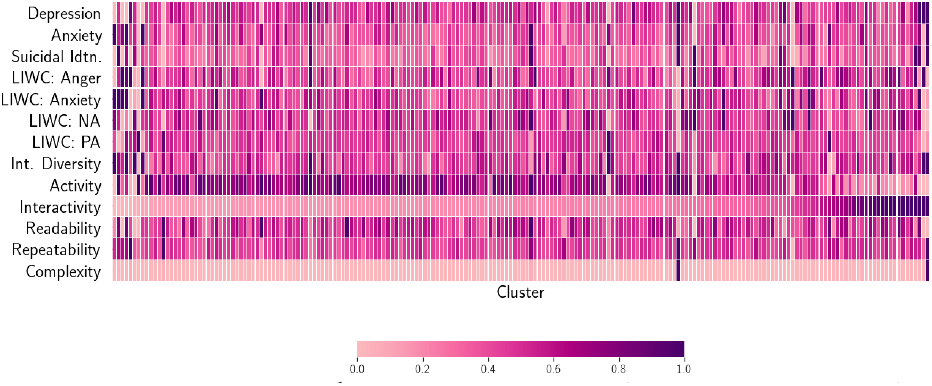
Heatmap showing mean values for a sample of covariates across 200 clusters. Values are rescaled using min-max scaling of 0 to 1 within a covariate.

#### Evaluating Balance

The purpose of matching is to ensure that confounders are minimized to the maximum extent due to individual differences, and to help conduct like-for-like comparisons. We evaluate the balance of the covariates using standardized mean differences (SMDs) across covariates in *Case* and *Control* groups. Two groups are considered to be balanced if the covariates reveal SMD lower than 0.25 (Kıcıman et al. 2018)— a condition not fulfilled in only 0.15% cases (364 out of 207,844 covariate-cluster combinations). Further, a significant drop (57%) in mean SMD from 0.16 (sd = 0.13) in the unmatched dataset to 0.07 (sd = 0.08) in the matched dataset, indicates a good balance (Figure 4c).

## 5 Potential Causes of Psychosocial Outcomes

We now study the factors that potentially contribute to psychosocial changes. Below we list potential treatment measures that are based on the literature as contributors to psychosocial change. We hypothesize that these factors, both implicit and explicit in the responses contribute to an individual’s psychosocial outcomes. Some of these factors are likely to be correlated among each other, aligning with the fact that “causes” of ones’ psychosocial outcomes are inherently coarse and complex combination of these factors.

### Number of Responses

We hypothesize that *Case individuals received greater number of responses to their concerns*. We rationalize that receiving more responses is associated with greater social support and sense of belonging in community, which have been found to be effective in an individual’s psychosocial improvement in psychology literature (Glass and Maddox 1992). To test this hypothesis, for every individual, we calculate the average number of responses received per post in the treatment phase, and compare these averages in the matched samples of *Case* and *Control* individuals.

### Verbosity

Along the lines of the above, we hypothesize that *Case individuals received more verbose or longer responses to their concerns*. We compare the length of responses in terms of the number of words and number of unique words received by the *Case* and *Control* individuals.

### Immediacy

Because immediate and sooner responses are generally recommended in the cases of mental health crisis (Flannery and Everly 2000), we hypothesize that *Case individuals received more immediate responses*. Essentially, we computed the average time to the first response received by the *Case* and *Control* individuals.

### Diversity/ Creativity

Drawing on the efficacy of counseling and psychotherapy styles (Althoff et al. 2016, Norcross and Lambert 2018), we hypothesize that *Case individuals received more diverse responses, in comparison to the Control individuals who received more templated and generic responses*. To examine this, we obtain the lexico-semantic diversity within the responses received by the *Case* and *Control* individuals. In particular, we use the 300-dimensional word embedding vector representations (Mikolov et al. 2013). For the responses in either of *Case* or *Control* corpus, we find their average cosine distance from the centroid of the corresponding corpus. This operationalizes the diversity in responses within *Case* and *Control* corpuses.

### Emotionality

We hypothesize that *Case individuals received responses that contained greater emotions and positive affirmations* (Norcross and Lambert 2018). For this, we use LIWC to obtain the normalized occurrences of affective keywords in the responses received by the *Case* and the *Control* individuals (Pennebaker, Mehl, and Niederhoffer 2003).

### Adaptability

We hypothesize that *Case individuals received responses that were more customized and attuned to their concerns*. This draws upon literature postulating that adaptable and linguistically accommodating responses are more effective in support than templated or generic responses (Althoff et al. 2016; De Choudhury and Kıcıman 2017). Better adaptability aids improved social feedback, solidarity, social exchanges, and reciprocated feelings of intimacy (Ferrara 1991). We examine adaptability in two measures, *topical congruence* and *linguistic style accommodation*.

#### Topical Congruence

Motivated by Pennebaker et al.’s (2003) work that content words are indicative of numerous psychosocial aspects, we extract the content words in responses and posts (using LIWC). We operationalize topical congruence between a response and the original post as the lexicosemantic similarity between the two, for which we obtain the cosine similarities between their word embedding representations (Das Swain et al. 2020; Tan et al. 2016).

#### Linguistic Style Accommodation

We obtain linguistic style accommodation of each query to by using Linguistic Style Matching (Sharma and De Choudhury 2018). We compute the cosine similarity of each response and original post on the normalized occurrences of non-content or linguistic style dimensions — these are function words across the categories of articles, prepositions, pronouns, auxiliary verbs, conjunctions, adverbs, negations, etc (Pennebaker et al. 2003).

### Credibility of the Responders

People tend to show trust in more credible and reputable individuals in the community (Ma et al. 2019). Accordingly, we hypothesize that *Case individuals received responses from those who are experienced “care and support” givers in the community*. To measure responders’ experience of providing support on the platform, we quantify their tenure (or duration of time spent), interactivity (ratio of number of responses to number of posts), and activity (number and rate of posting) on TalkLife.

### Language Style of Responses

Literature posits the importance of language style in effective psychotherapy (Norcross and Lambert 2018). Using personal opinion induces a sense of belonging, and also corresponds to mindful genuineness on the part of the peer-supporter. The nature of communication is a direct correlate of the complexity of language (Kolden et al. 2011). Language style can be characterized as categorical and dynamic (Pennebaker et al. 2014). Theoretically, categorical language includes approaching the world in a relatively logical, complex, and “amateur scientist” manner, and dynamic language is typically used by individuals who are more socially engaged, tell stories, and pay more attention to the world around them. We hypothesize that *“the responses received by the Case individuals is more dynamic”*. We adopt the measure of categorical-dynamic index (`CDI`) proposed by Pennebaker et al. (2014). This is a bipolar index, where higher `CDI` indicates a categorical style, and lower `CDI` indicates a dynamic or narrative style. In particular, `CDI` for a given text is quantified based on the percentage of words per style related parts of speech:

> *`CDI` = (30 + article + preposition – personal pronoun – impersonal pronoun – aux. verb – conjunction – adverb – negation)*

### Nature of Support

Past work suggests that social support is greatly effective in helping individuals cope with mental health struggles (Kummervold et al. 2002). Situated in the “Social Support Behavioral Code”, two forms of support that have received theoretical and empirical attention are *emotional* and *informational* support. Emotional support corresponds to language containing empathy, encouragement, and kindness, and is considered to be most effective form of psychosocial support (Sharma and De Choudhury 2018).

Informational support, corresponds to providing information and advice, and is also known to be effective in positively impacting perceived empathy (Nambisan 2011). We hypothesize that *Case individuals received greater support*.

We obtain the presence of emotional (ES) and informational (IS) support in responses. We use a dataset built and expert-verified in prior work (Sharma and De Choudhury 2018) that labels supportive responses on Reddit on the degree of ES and IS. We build two binary SVM classifiers with linear kernel, either of which characterizes the degree (high/low) of ES or IS in a post. The *k*-fold cross validation accuracy (*k* = 10) of ES and IS classifiers are 0.71 and 0.77 respectively (Table 2 reports accuracy metrics, Figure 6 shows ROC curves). While the classifiers are expected to perform well, better accuracy can be achieved with sophisticated models and expert annotation on TalkLife, our objective here is to leverage the feasibility of measuring nature of support in language. Note that similar to the mental health classifiers, support classifiers are also transferred from Reddit to TalkLife data, and the linguistic similarity between the two datasets ensures a reliable transfer (Section 4). We use these classifiers to machine label all responses — 15% responses contain ES and 3.3% responses contain IS, and then compare their prevalence per *Case* and *Control* group.

**Table 2:**
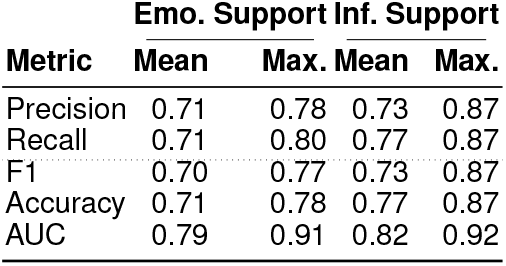
Accuracy metrics of the Support Classifiers.

**Figure 6:**
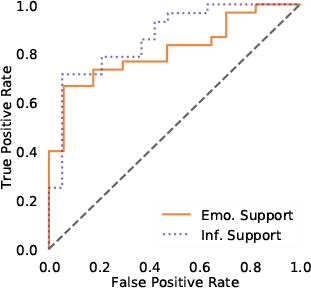
ROC Curve of Support Classifiers

## 6 The Effect of Supportive Interventions

Per previous section, we now test the hypotheses to quantify the differences per treatment across the matched samples of *Case* and *Control* individuals. We obtain effect size (Cohen’s *d*), and evaluate statistical significance in differences using independent sample *t*-test. We conduct Kolmogorov-Smirnov (*KS*) test which essentially tests against the null hypothesis that the distributions of treatments in the *Case* and *Control* groups are drawn from the same distribution. Table 3 summarizes these differences, which we discuss here.

**Table 3:** Summary of differences in responses received by *Case* and *Control* individuals. We report average occurrences across matched clusters, effect size (Cohen’s *d*), independent sample *t*-statistic, and *KS*-statistic. Rows with significant differences are shaded in grey, *p*-values are reported after Bonferroni correction (* *p<*0.05, ** *p<*0.01, *** *p<*0.001).

| Measure | Case | Control | $d$ | $t$ | $KS$ |
| --- | --- | --- | --- | --- | --- |
| Num. Responses | 16.34 | 12.98 | 0.09 | 0.88 | 0.13 |
| Verbosity (Avg. Per Response) |  |  |  |  |  |
| Num. Words | 19.98 | 18.07 | 0.96 | 9.08*** | 0.43*** |
| Num. Unique Words | 17.60 | 16.19 | 0.97 | 9.20*** | 0.48*** |
| Immediacy (Minutes) | 6.22 | 5.95 | 0.11 | 1.05 | 0.09 |
| Diversity/Creativity | 0.66 | 0.63 | 1.09 | 9.18*** | 0.46*** |
| Emotionality (% Words) |  |  |  |  |  |
| Anger | 0.71 | 0.70 | 0.05 | 0.43 | 0.16* |
| Anxiety | 0.33 | 0.31 | 0.15 | 1.41 | 0.12 |
| Sadness | 0.48 | 0.47 | 0.08 | 0.73 | 0.13 |
| Neg. Affect | 0.87 | 0.87 | 0.00 | 0.02 | 0.11 |
| Pos. Affect | 5.67 | 5.49 | 0.21 | 1.91** | 0.12** |
| Swear | 0.36 | 0.36 | 0.04 | 0.38 | 0.17* |
| Adaptability |  |  |  |  |  |
| Topical Congruence | 0.65 | 0.61 | 1.22 | 11.55*** | 0.50*** |
| Linguistic Accommodation | 0.80 | 0.61 | 1.38 | 18.14*** | 0.63*** |
| Credibility of Responders |  |  |  |  |  |
| Tenure (days) | 233.93 | 234.49 | -0.00 | -0.06 | 0.08 |
| Interactivity | 22.46 | 19.39 | 0.32 | 2.93** | 0.13* |
| Num. Posts | 2987.34 | 2721.91 | 0.61 | 5.75*** | 0.40*** |
| Posts per Day | 11.73 | 11.01 | 0.15 | 1.42 | 0.10 |
| Language Style (CDI) | 3.39 | 4.10 | -0.45 | -3.74*** | 0.25*** |
| Nature of Support |  |  |  |  |  |
| Emotional | 0.20 | 0.17 | 0.93 | 8.82*** | 0.36*** |
| Informational | 0.05 | 0.04 | 0.55 | 5.20*** | 0.31*** |

### Number of Responses

Figure 3d shows the distribution of number of responses received by *Case* and *Control* individuals in the treatment period. While on an average, *Case* individuals receive 25% more responses than their matched *Control* individuals, we find no significant differences in the number of responses received by the matched *Case* and *Control* individuals. This suggests that individuals with similar concerns, and psychological and social attributes (because of the matching framework), are likely to receive similar number of responses. Therefore, we cannot reject the null hypothesis that the number of responses received by *Case* and *Control* individuals are from the same distribution.

### Verbosity

*Case* individuals receive more verbose responses than the matched *Control* individuals. This is revealed by both average length of response (*t* = 9.08, *p<*0.05), and average number of unique words per response (*t* = 8.91, *p<*0.05), where *Case* individuals receive 11% more words, and 9% more unique words per response. This supports our hypothesis on the differences in verbosity, suggesting longer responses and lower repeatability of words are more likely to help psychosocial improvement.

### Immediacy

We find no significant difference in immediacy or the time to respond to posts. This could be because we study long-term and averaged-out improvements of psychosocial outcomes, rather than short-term bursts. Again, platform-specific design and post ranking on homepage plausibly does not distinguish the type and criticality of concern leading to all posts being responded back in similar intervals of time. This phenomenon is further revealed by the low standard deviation (*∼*12 mins.) in the time-to-first-response across all the responses in our TalkLife dataset. That said, immediacy is considered to be essential for coping with critical and crisis circumstances, and this leaves room for future investigations on the prevalence of such instances on TalkLife.

### Diversity/ Creativity

Supporting our next hypothesis, we find that the responses received by *Case* individuals are typically more diverse. The average distance (or diversity) among the responses received by the *Case* individuals is 5% higher (*t* = 9.18, *p* < 0.05). This also hints at the possibility that the *Control* individuals received more generic and templated responses. To understand this better in context, we inspect a few top keywords in responses received by the *Case* and *Control* users, to find many generic responses such as *hope great day, wish good luck*, etc. in responses to *Control* individuals.

### Emotionality

As Table 3 indicates, we find significant differences in the expression of positive affect — *Case* individuals received 3.5% greater positive affect. This aligns with literature that greater positivity is associated with effective psychotherapy (Truax and Carkhuff 2007). In contrast, we find no significant differences in emotionality across anger, anxiety, sadness, negative affect, and swear. Nonetheless, a common trend across all the affective attributes is that the responses received by the *Case* individuals show a greater occurrence than that by *Control* individuals. Together, our hypothesis on emotionality is only partially supported.

### Adaptability

We measure adaptability of the responses in terms of topical congruence and linguistic style accommodation. Topical congruence occurs 6.6% higher in the responses received by the *Case* individuals than the *Control* individuals (*t* = 11.55, *p<*0.05). In terms of linguistic accommodation, the responses received by the *Case* individuals show 31.15% greater (*t* = 18.14, *p<*0.05) linguistic style matching than the ones received by the *Control* individuals. Both the measures of adaptability, therefore, support our hypothesis, aligning with prior work that greater adaptability in responses is associated with increased supportive outcomes.

### Credibility of the Responders

To examine if responder credibility significantly varied between *Case* and *Control* responses, we measure the differences in the responders’ tenure (number of days on the platform), interactivity, number of posts, and the frequency of posting behavior (posts per day). Among these, we find no significant differences in the tenure and the number of posts per day. However, we find 16% greater interactivity and 10% greater number of posts for the responders to *Case* individuals as compared to that to the *Control* individuals. This suggests that responses from members who are more active on the platform seem to be typically more effective. Drawing on prior work, it may be associated with the fact that the members who are more experienced with the platform use more linguistically accommodating language or probably learn over time in what constitutes more supportive responses. Supporting our hypothesis, we find that *Case* individuals greatly received responses from those who are experienced “care and support” givers in the community.

### Language Style of Responses

We find that the average Categorical Dynamic Index (`CDI`) of responses received by *Case* individuals is 17% lower. This suggests that the responses received by *Case* are more dynamic in nature, or exhibit a dynamic style of thinking including a focus on others (such as greater use of pronouns), time-based stories, and use of simpler words (Pennebaker et al. 2014). Supporting our hypothesis, we conjecture that dynamic style of writing is likely to help psychosocial improvements on the platform.

### Nature of Support

We find that responses to *Case* is higher in both emotional and informational support. Among these, *Case* individuals receive 18% greater emotional support, and 25% greater informational support. Therefore, our hypothesis is supported, and we conjecture that greater support contributes to better psychosocial improvement on the platform. While prior work (De Choudhury and Kıcıman 2017) compared the efficacy of emotional against informational support, our work finds that both kinds of support are effective towards psychosocial improvement.

### Summary

We find that many of the treatment measures positively impact long-term psychosocial outcomes. We can use Cohen’s *d* to rank treatments by their efficacy. Based on *Case* and *Control* means from Table 3, we can construct a binary treatment with a mean-split threshold on *Case* and *Control* means. This maybe interpreted as treatments with high Cohen’s *d* will likely ensure higher fraction of (binary) treated individuals with outcomes similar to the *Case* group. Our results indicate adaptability, diversity, verbosity, and emotional support rank highest in differentiating *Case* and *Control* individuals and thus can be considered as preferable treatment candidates for a future randomized experiment.

### Robustness of Findings

Recall that our study design relies on chosen values of *n*_1_ and *n*_2_ posts to define pre-treatment, treatment, and post-treatment phases (Figure 2). We test if our findings hold robust for a variety of (*n*_1_, *n*_2_) combination pairs. For different pairs, we re-conduct our entire study including measuring outcomes, conducting matching, testing balance, and computing differences in treatment for matched *Case* and *Control*. Figure 7 shows the differences as effect-size (Cohen’s *d*) across (*n*_1_,*n*_2_) pairs of (3,8), (2,6), (3,5), (4,4), (5,3), (6,2), (3,6), and (4,6). We find that effect size is very similar across all combinations of (*n*_1_, *n*_2_), showing a low standard deviation of 0.08 on average. Again, the treatment measures consistently show similar statistical significance as per *t*-test and *KS*-test. All these significant measures also show the *same* directionality of differences, such as verbosity, diversity, and adaptability are uniformly greater, and `CDI` is uniformly smaller for responses received by *Case* individuals.

**Figure 7:**
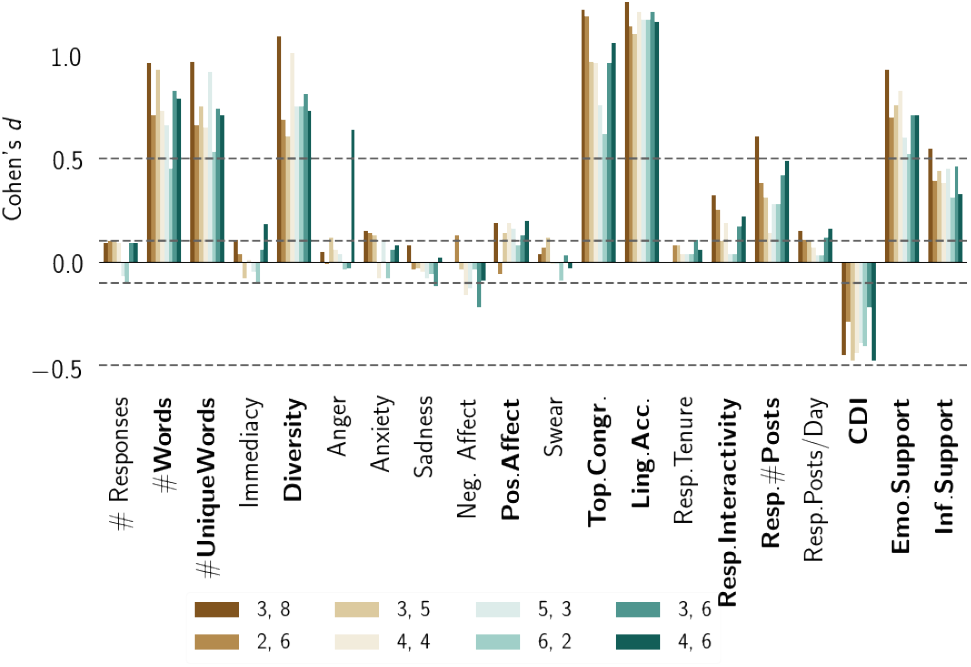
Cohen’s *d* of treatment differences in responses received by *Case* and *Control* users for several combinations of (*n*_1_, *n*_2_) breakpoints defining pre- and post-treatment phases. Boldfaced measures show statistical significance in *t*-test and *KS*-test (*p* < 0.05). Statistically significant measures show consistent directionality in differences.

Another component of our work concerns the decision to separately estimate the treatment differences between Case and Control for each treatment, rather than considering their effects together and also including any interaction effects. While we measure the differences for our theory-driven treatments independently, these treatment measures can be correlated and interdependent, e.g., positive affect and emotional support. Our study design is motivated towards providing interpretative understanding of how different psychotherapeutic measures function towards psychosocial improvement in OMHCs. Still, to test the robustness of such independent comparisons, we construct regression models with treatment measures as independent variables and overall psychosocial outcome as (continuous) dependent variable. We control our models with the same covariates used in matching. We eliminate correlated features using variance inflation factor (threshold = 10) (Das Swain et al. 2019, Miles 2014). We also include regularizations (L1, L2) and interaction terms (degree 2) in the regression models. We find that all the interaction terms show statistically insignificant effects. The regularized and unregularized models show similar coefficients. For linear interpretability, Table 4 reports the unregularized model’s coefficients of treatment measures found to be significant. The magnitude of regression coefficients is interesting and inspires further theoretical and empirical investigations. Consistent with our previous analysis, we find that the directionality of the regression coefficients agree with that we found by independently testing the treatment measures (Table 3).

**Table 4:** Coefficients of linear regression of treatment measures as independent variables and psychosocial outcomes as dependent variables. Only statistically significant coefficients are reported (* *p<*0.05, ** *p<*.01, *** *p<*0.001).

| Measure | Coefficient | Measure | Coefficient |
| --- | --- | --- | --- |
| Num. Words | 7.32e - 5*** | Topical Congruence | 2.11e - 2*** |
| Diversity | 1.03e - 6** | Resp. Interactivity | 2.01e - 5* |
| Emotional Support | 5.55e - 3*** | CDI | -3.52e - 5** |
| Informational Support | 8.33e - 4* | Anger | -4.72e - 3* |

The consistency of results via different approaches confirm that our findings are robust and not sensitive to choice of treatment periods or specific estimation methods, but rather a reflection of the phenomenon in our context of TalkLife.

## 7 Discussion and Conclusion

We studied factors that contribute to psychosocial changes in online mental health communities (OMHCs) using a case-control design. We examined whether effective support factors identified in psychotherapy literature are also similarly effective in OMHCs. Confirming past work, we find that factors such as diversity, adaptability, positivity, supportive nature, and dynamicity of language in responses are positively associated with effective psychosocial support. In contrast, simple factors such as immediacy and quantity of responses show insignificant effects on psychosocial outcomes. Our findings can be used to rank potential interventions for peer supporters. We discuss these points below.

### Methodological implications

Our work provides a useful alternative to cohort-based analyses for studying cause-and-effect in online communities, especially when the outcome is well-specified and treatments are continuous variables. In such cases, our approach can help study two dimensional changes in the treatments — 1) along the breadth, that is studying several treatments together, and 2) along the depth, that is how much of a treatment is necessary.

From a treatment dosage perspective, each measure considered in our study is a continuous variable, and it is often not possible to determine an appropriate binary cutoff for (no) treatment in a prospective causal-inference setup. Our approach avoids this limitation by focusing on the differences in the treatment measures in *Case* and *Control* groups — and suggesting the dosage of measure required for desirable outcomes across a subpopulation (Table 3). This can be useful for design interventions on a social computing platform. For instance, TalkLife can propose guidelines that recommend expectations to the members in what ways they would be helped. Also, such differences can be used to formulate treatment cutoff in conducting careful experimental studies to verify and adopt design changes. More generally, our approach allows examining several treatments that potentially contribute to the same desirable outcome. Because the effects of each treatment can vary across individuals, such a study design helps to identify which treatment or combinations of treatments could be effective for certain individuals.

### Implications for OMHCs

Implications for OMHCs. Given that OMHCs largely rely on amateur peer supporters, one of the biggest questions is how to help supporters write more effective responses. By comparing factors that lead to a positive outcome, our work provides evidence on effective support factors. We provide a way to rank and compare potential interventions so that effective candidate treatments can be considered and encouraged. For example, based on our results, OMHCs may nudge members to write more positive or adaptive responses. Our work also contributes to digital therapeutics, given limited availability of trained psychotherapy providers, we believe insights drawn from our work can be useful to train peer-supporters and volunteers who want to help in OMHCs (Kazdin 2011; Torous and Hsin 2018). That said, causal evidence from observational studies comes with the assumption that all confounders were conditioned. As randomized experiments are the gold standard to measure efficacy, our work provides a means to prioritize which treatments to consider for such experiments of understanding effective interventions.

### Towards personalized support

From the perspective of individualized and precision medicine, our work builds the case for patient-centered and personalized psychotherapeutic care (Shippee et al. 2012). We find that, just like in face-to-face settings, templated and generic responses are not as effective as personalized and adaptive responses. Applying our approach of stratifying (or clustering) individuals based on psycholinguistic and psychosocial similarity may enable decision-making on what combination of treatments can be more effective in particular clusters. This can help design frameworks to tailor treatments per cluster of individuals.

### Ethical Implications

Despite the potential, there are important ethical implications associated with using such quantitative analyses in practice. Privacy considerations should be made when machine guided interventions are tailored to OMHC participation. We expect analyses to be over deidentified datasets and interventions to be restricted to the online platform, ensuring that such analyses cannot be used to monitor one’s trajectory of mental health and make offline decisions based on it. There are potential civil and ethical liability concerns in providing machine-guided support in an online medium, leaving room for further discussions on adopting these approaches in practice (Chancellor et al. 2019).

### Limitations and Future Work

Our work has limitations, which also suggest promising future directions. We do not account for spill-over and passive engagement effects, e.g., individuals may be helped by browsing discussion threads of mental health support. We only consider mean-aggregated psychosocial outcomes, which is likely unable to capture shorter changes of psychosocial outcomes, e.g., individuals who show show fluctuating affective states or mood instability, which maybe accounted for by using complementary data sources (Morshed et al. 2019). Our operationalization does not unpack intricacies in each psychosocial outcome separately, and does not encompass all mental health conditions; certain psychosocial changes (e.g., activity) considered to be positive in our study may not be applicable in certain mental health conditions (e.g., ADHD). Future work can address these concerns by examining psychosocial outcomes per condition, in a fine-grained temporal fashion.

Because our work examines observable behavior on Talk-Life and plausibly excludes offline and latent individual differences, we cannot establish clinical validity. Future work obtain consented data (Saha et al. 2019) and expert-appraisal (Ernala et al. 2018, Levonian et al. 2020) to validate our findings with greater rigor. We only study those who show continued participation on the platform. This “dropouts” issue is also encountered in experimental and randomized trial settings (Lindsey 2000), and future work can include measures like likelihood of failed interactions (Zhang et al. 2018) and survival analysis to incorporate the behavior and dropping out of participants from platform (Ma et al. 2017, Yang et al. 2017). For treatments, we can examine the effects of social ties, social capital (Burke et al. 2010), cross-cultural accommodation (Pendse et al. 2019), and other linguistic attributes, such as politeness (Zhang et al. 2018), stance (Pavalanathan et al. 2017), and brevity (Gligorić et al. 2019).

As any other observational study, we recognize that we do not infer “true causality”. We cannot eliminate the likelihood of type II errors, a vulnerability of retrospective causal design. Gelman and Rubins (2013) argue that reverse-causal problems are better studied with forward-causal questions, and Watts (2014) notes the impossibility to test all explanations simultaneously. While acknowledging these concerns, we believe our work is a step towards understanding the effects of a variety of heterogeneous factors in psychosocial outcomes. Accounting for all possible confounds is technically infeasible, and our work only *minimizes* the confounds by using a variety of theory-driven covariates, thereby providing insights beyond simpler correlational analyses. Alternative study designs such as instrumental variable methods may help to further minimize confounding biases. While our study only considers a finite set of treatment measures, more measures can be easily plugged in to understand their effectiveness. Our study design facilitates a simple but robust mechanism to understand the factors associated with psychosocial outcomes in an online setting, and in turn helps us draw actionable insights and implications towards running confirmatory randomized experiments and designing effective OMHCs.

## Data Availability

This paper uses sourced data (licensed and consented) from TalkLife. The raw data will not be publicly shared for privacy and ethical reasons given the sensitivity of the work.

## 8 Acknowledgement

Saha conducted this work while at Microsoft. We thank TalkLife for their support. We thank Monojit Choudhury, Munmun De Choudhury, Daejin Choi, Sindhu Ernala, Emre Kıcıman, Vedant Das Swain, Taisa Kushner, Sachin Pendse, Ian Stewart, Adith Swaminathan, and Dong Whi Yoo for their help and feedback.

